# Parent-of-origin effects in the life-course evolution of cardio-metabolic traits

**DOI:** 10.1101/2021.10.28.21265599

**Authors:** Rucha Wagh, Pooja Kunte, Chittaranjan S Yajnik, Rashmi B Prasad

## Abstract

**Objective:** Human traits are heritable, and some of these including metabolic and lipid phenotypes show preferential parental transmissions, or parent-of-origin effects. These have been mostly studied in populations comprising adults. We aimed to investigate heritability and parent-of-origin effects on cardiometabolic and anthropometric traits in a birth-cohort with serial measurements to assess if these effects manifested at an early age.

**Research design and methods:** We investigated heritability and parent-of-origin effects on cardiometabolic and anthropometric traits in the Pune Maternal Nutrition Study (PMNS) wherein offspring and parents were studied from birth and followed-up for 18 years. Heritability was estimated by calculating association between mid-parental phenotypes and offspring. Maternal and paternal effects on offspring phenotype were modelled by regression after adjusting for age, sex and BMI. Parent-of-origin effects were calculated by the difference between maternal and paternal effects.

**Results:** Anthropomorphic traits and cardiometabolic traits were robustly heritable. Parent-of-origin effects were observed for glycemic traits at both 6- and 12-years, with a paternal effect at 6-years which transitioned to a maternal effect at 12-years. For insulin and HOMA-S, a negative maternal effect transitioned to a positive one at 12-years. For HOMA-B, a paternal effect at 6-years transitioned to a maternal one at 12-years. Lipid traits consistently showed stronger maternal influence while anthropometric traits did not show any parental biases.

**Conclusions:** Our study highlights that parental programming of cardiometabolic traits is evident from early childhood and can transition during puberty. Further studies are needed to determine the mechanisms of underlying such effects.

## Introduction

Human traits and diseases are a consequence of a complex interplay between genetics and environment. Assessment of heritability provides information on the contribution of the genetic component to the total phenotypic variation in a population. Anthropometric and metabolic traits have thus far been shown to be heritable to varying degrees (1; 2). Genetic association studies have identified a number of variants associated with these traits, however, the proportion of heritability attributed to these variants was rather limited (3; 4).

In a classic Mendelian pattern of transmission, a trait can be contributed by both the parents equally; it is also possible that these may be inherited preferentially from one of the parents while the contribution of the other parent can be low, neutral or even opposite. Such effects whereby the expression of the phenotype in the offspring depends upon which parent they are inherited from, are termed as parent-of-origin effects. These can be attributed to genetic imprinting, intrauterine effects, or maternally inherited mitochondrial genes (5). The significance of such effects in aetiology of type 2 diabetes and obesity has been emphasized previously (6). Type 2 diabetes shows a preferential maternal transmission (2; 7), and a substantial component may originate in the intrauterine period. Several studies have demonstrated that early life exposures can influence developmental programming and increase risk to cardiometabolic disorders in later life (8-10).

Parent-of-origin as well as sex-specific parental effects were observed for anthropometric measures, insulin secretion and all cholesterol levels (1; 2; 11). For instance, sons of diabetic mothers had lower insulin concentrations compared to those of diabetic fathers, while daughters of diabetic mothers had the lowest high-density lipoprotein (HDL) levels (2).

These studies were of cross-sectional design in adult offspring; however, given the role of early life programming in risk of cardiometabolic disorders in later life, it is possible that these parental effects manifest at an early age. The Pune Maternal Nutrition Study (PMNS), a well-characterised prospective birth cohort provides a unique opportunity to study heritability and parent-of-origin effects in parent-offspring trios in a life-course model. In this study, we investigated the heritability and parent-of-origin effects of anthropometric, glycemic and insulin related traits and lipid traits in the PMNS birth cohort with follow-up from birth through puberty till adulthood. Parent-offspring associations and transitions of parental specific effects across childhood was assessed.

## Methods

### Cohort characteristics

The Pune Maternal Nutrition Study (PMNS) (Figure 1) was established in 1993 in six villages near Pune, to prospectively study associations of maternal nutritional status with fetal growth and later diabetes risk in the offspring. Married, non-pregnant women (N=2,466) were followed up. Between 1994 and 1996, those who became pregnant (F0 generation) were recruited into the study. The children’s (F1 generation) growth was measured *in utero*, at birth and 6 monthly thereafter, and body composition and glucose-insulin indices were measured 6-yearly (Table 1, Supplementary Table 1). Ethical permission and informed consent were obtained for the study. The study was approved by village leaders and the KEM Hospital Research Centre Ethics Committee. Parents gave written consent; children under 18 years of age gave written assent, and written consent after reaching 18 years.

**Figure 1.**
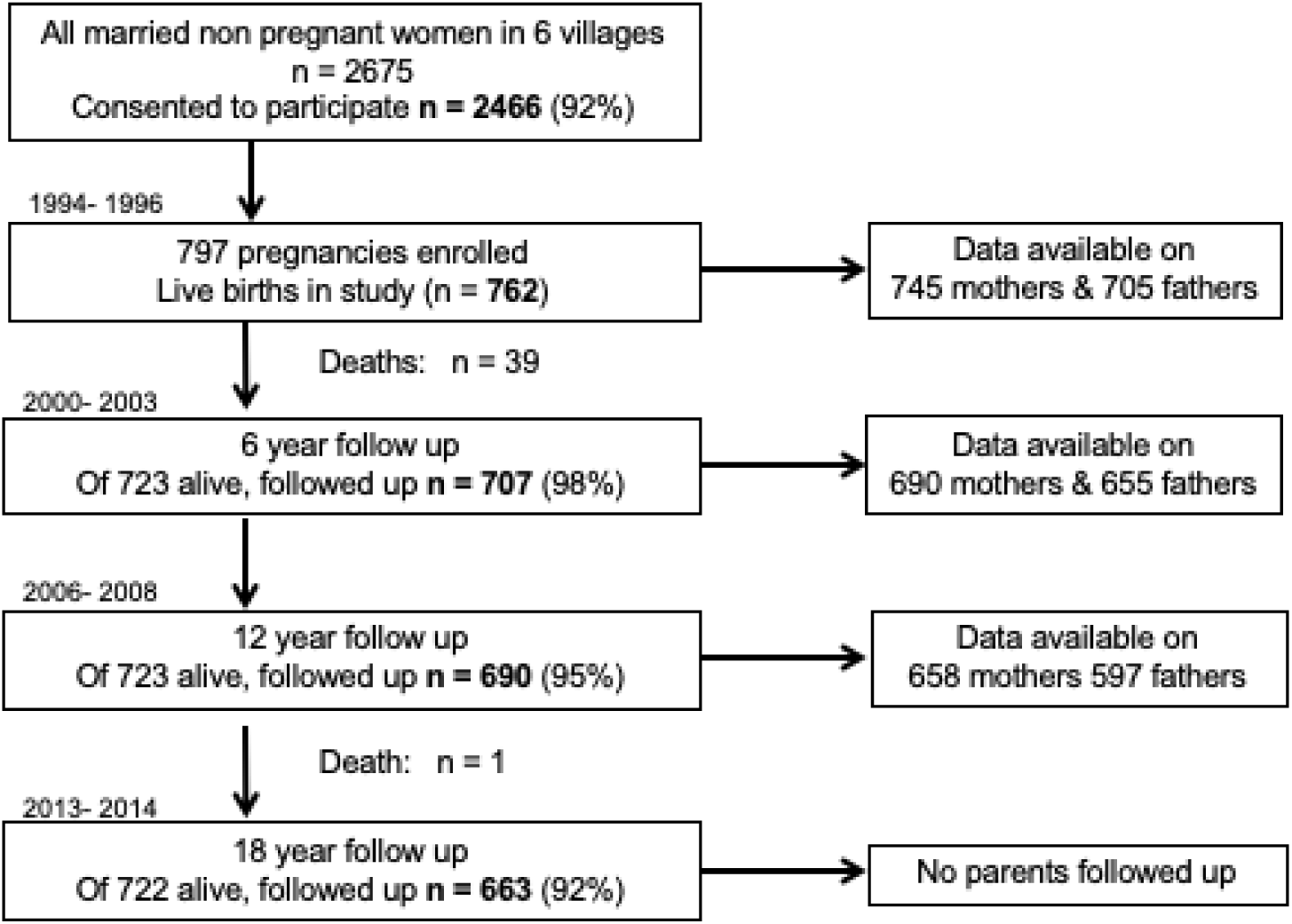
STROBE flow diagram of the Pune Maternal Nutrition Study. *\* Maximum numbers available are mentioned. Not all data may be available on the mentioned numbers*.

**Table 1.**
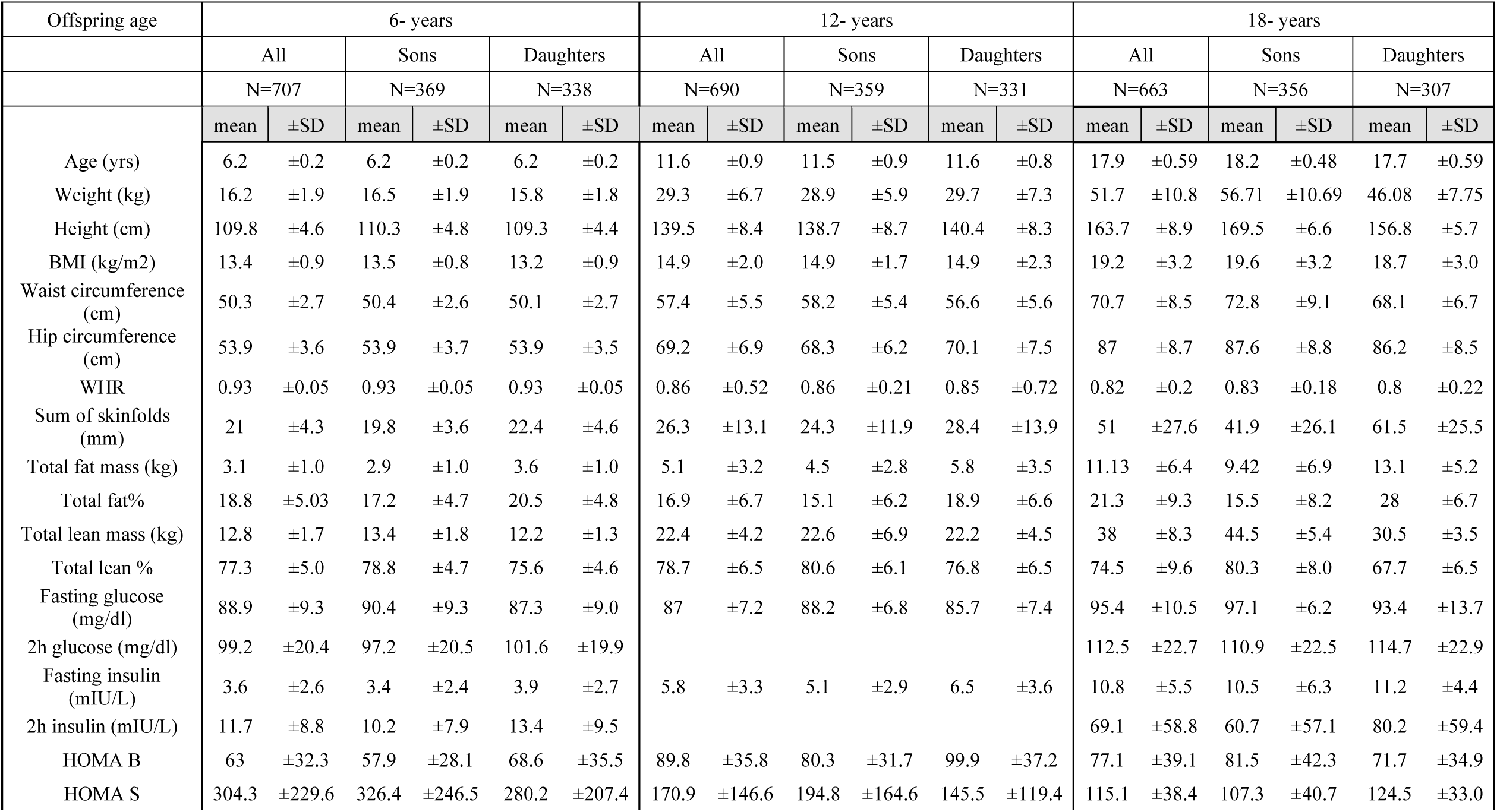
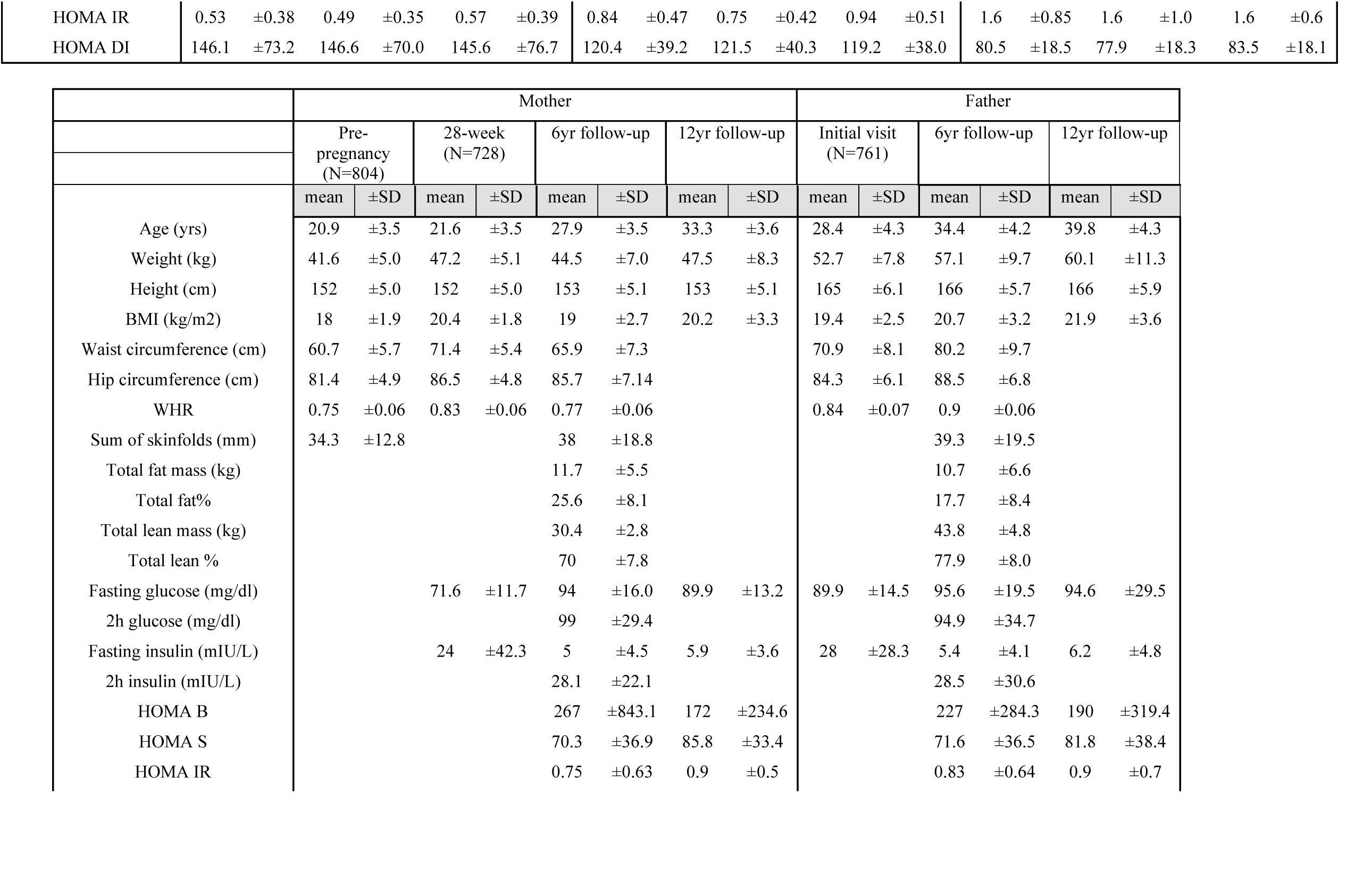
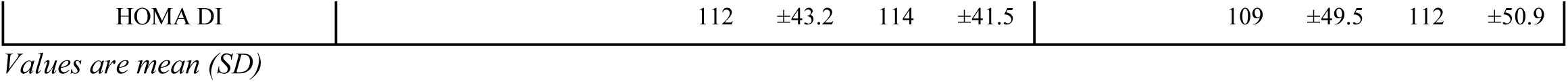
Description ofoffspring and parents of the Pune Maternal Nutrition Study (PMNS) for visits at offspring ages 6-, 12- and 18- years.

### Anthropometric and clinical measurements in parents and offspring

Newborn anthropometry including weight, length, abdominal circumference and skinfolds was carried out within 72 hours of birth.

Comprehensive assessments of body composition and glucose and insulin concentrations were made at 6, 12 and 18 years. Participants arrived at the Diabetes Unit (KEM Hospital, Pune) the evening before, had a standardized dinner, and fasted overnight. In the morning, a fasting blood sample was collected. At 6 years, an oral glucose tolerance test (OGTT) was performed, using 1.75g/kg of anhydrous glucose, followed by further samples at 30 and 120 minutes. At 12 years, only a fasting sample was collected. At 18 years a full OGTT (75g anhydrous glucose) was repeated.

Glucose was measured by the glucose oxidase/peroxidase method, and specific insulin by ELISA (Supplementary Table 2). Homeostatic model assessment for insulin sensitivity (HOMA-S), beta-cell function (HOMA-ß) and insulin resistance (HOMA-R) were calculated using data from the fasting samples and the iHOMA2 website ((https://www.phc.ox.ac.uk/research/technology-outputs/ihoma2) (12). Disposition index (β cell function adjusted for insulin sensitivity) was calculated as HOMA-S*HOMA-β.

Total fat and lean mass and body fat% were measured by Dual Energy X-ray Absorptiometry scanner. (Lunar DPX-IQ 240 pencil beam machine, Lunar Corporation, Madison, WI, USA). Body size (anthropometry) and glucose tolerance (75 g OGTT) were measured in both parents at the time of initial visit and at 6 year follow up. Body size and only a fasting blood test was available at 12-year follow-up. BMI was calculated using standard formula [weight (kg)/square of height (m)] and WHR was calculated as waist circumference (cm) / hip circumference (cm).

### Statistical analysis

Heritability and parent-of-origin effects were assessed between F0 and F1 generation across different time points (Pre-pregnancy, At birth, 6yrs, 12yrs, 18yrs) in PMNS cohort for anthropometric and glycemic traits. For this, the skewed variables were log transformed, and all variables were standardized (mean zero, standard deviation unity) adjusted for age and gender to facilitate the comparison between variables. Heritability was estimated using regression models adjusted for age and gender expressed in ß and p-value. Parent-of-origin effects were tested by computing the difference in maternal and paternal regression coefficients using the formula [(b1-b2) / sqrt (seb1**2 + seb2**2 – cov(b1*b2))] expressed in Z and corresponding p value.

## Results

### Heritability of anthropometric and metabolic traits

To investigate the proportion of offspring phenotypic trait attributable to parent phenotype variation, we calculated heritability estimates of anthropometric and metabolic traits measured in ∼700 parent-offspring trios in the PMNS cohort at birth, 6, 12 and 18 years of age (Table 1). Offspring’s weight, height and BMI were significantly associated with corresponding mid-parental measures with coefficients ranging between 0.13 to 0.24 at birth. Moreover, there was an upward trend in effect size across timepoints increasing to 0.36 to 0.51 at 6-years, 0.44 to 0.52 at 12-years, and 0.41 to 0.60 at 18-years (Supplementary Table 3). Offspring measurements of waist and hip circumference, WHR, fat and lean mass (DXA) at 6-years also showed significant association with mid-parental measures. These measurements were available in parents only at 6y (Supplementary Table 3). These heritability estimates were similar for sons and daughters (Supplementary Table 3).

Offspring concentrations of fasting glucose and insulin, as well as HOMA 2B showed a significant association with corresponding mid-parental measures at 6 years (ranging from 0.10 to 0.41). At 12-years, similar associations were observed for the aforementioned measures with addition of HOMA2S (ranging from 0.19 to 0.31). This trend was consistent for sons and daughters (Supplementary Table 4).

Triglyceride, total cholesterol and HDL levels were significantly heritable at 6 and 12 years. Considered separately, similar associations were observed in both sons and daughters with an exception of HDL levels at 12 years in daughters (Supplementary Table 4).

### Parent-of-origin and sex-specific parental effects on cardiometabolic traits

We next examined if there was a stronger association between the trait of the offspring and the trait of each of the parents specifically. If offspring showed a stronger association with the mother’s traits compared to the father’s, this would indicate a maternal effect and likewise for the paternal effect. If there were a significant difference between the maternal and paternal effects, this is indicative of the parent-of-origin effect.

### Anthropometry

While offspring anthropometry was significantly associated with that of each of the parents, no significant parent-of-origin effects were observed either in all offspring or for sons and daughters separately (Table 2, Supplementary Table 5).

**Table 2.** Parent-of-origin test for offspring anthropometry. Association between offspring anthropometry measures with that of each of the parents are presented as beta values and se. Differences between maternal and paternal effects are presented as Z scores and p-values.

|  | At birth |  |  |  |  |
| --- | --- | --- | --- | --- | --- |
| All offspring | MC (N=745)<br>beta (SE) | P-value | FC (N=705)<br>beta (SE) | P-value | Z (P) |
| Weight (kg) | 0.157 (0.044) | 0.0001* | 0.081 (0.043) | 0.065 | 1.24 (0.22) |
| Height (cm) | 0.176 (0.042) | 0.0001* | 0.149 (0.043) | 0.0001* | 0.45 (0.65) |
| BMI (kg/m2) | 0.103 (0.045) | 0.019 | 0.093 (0.044) | 0.035 | 0.16 (0.87) |
| Sons | MC (N=386) |  | FC (N=370) |  | Z (P) |
| Weight (kg) | 0.131 (0.058) | 0.033 | 0.059 (0.056) | 0.337 | 1.32 (0.19) |
| Height (cm) | 0.165 (0.058) | 0.005* | 0.138 (0.056) | 0.017 | 0.33 (0.74) |
| BMI (kg/m2) | 0.109 (0.061) | 0.074 | 0.06 (0.060) | 0.322 | 0.57 (0.57) |
| Daughters | MC (N=353) |  | FC (N=342) |  | Z (P) |
| Weight (kg) | 0.196 (0.069) | 0.002* | 0.115 (0.068) | 0.068 | 0.52 (0.61) |
| Height (cm) | 0.188 (0.063) | 0.002* | 0.166 (0.068) | 0.007 | 0.24 (0.84) |
| BMI (kg/m2) | 0.102 (0.069) | 0.112 | 0.128 (0.066) | 0.046 | -0.27 (0.78) |
|  | 6-year |  |  |  |  |
| All offspring | MC (N=690)<br>beta (SE) | P-value | FC (N=655)<br>beta (SE) | P-value | Z (P) |
| Weight (kg) | 0.277 (0.037) | 0.0001* | 0.301 (0.037) | 0.0001* | -0.46 (0.65) |
| Height (cm) | 0.349 (0.034) | 0.0001* | 0.316 (0.035) | 0.0001* | 0.68 (0.50) |
| BMI (kg/m2) | 0.213 (0.038) | 0.0001* | 0.254 (0.038) | 0.0001* | -0.76 (0.44) |
| Sons | MC (N=363) |  | FC (N=342) |  | Z (P) |
| Weight (kg) | 0.276 (0.048) | 0.0001* | 0.319 (0.049) | 0.0001* | 0.45 (0.65) |
| Height (cm) | 0.373 (0.049) | 0.0001* | 0.299 (0.048) | 0.0001* | 1.08 (0.28) |
| BMI (kg/m2) | 0.208 (0.052) | 0.0001* | 0.230 (0.053) | 0.0001* | -0.30 (0.77) |
| Daughters | MC (N=327) |  | FC (N=313) |  | Z (P) |
| Weight (kg) | 0.280 (0.058) | 0.0001* | 0.282 (0.058) | 0.0001* | -0.51 (0.61) |
| Height (cm) | 0.322 (0.051) | 0.0001* | 0.339 (0.055) | 0.0001* | -1.81 (0.07) |
| BMI (kg/m2) | 0.220 (0.056) | 0.0001* | 0.277 (0.055) | 0.0001* | -0.73 (0.47) |
|  | 12-year |  |  |  |  |
| All offspring | MC (N=658)<br>beta (SE) | P-value | FC (N=597)<br>beta (SE) | P-value | Z (P) |
| Weight (kg) | 0.298 (0.038) | 0.0001* | 0.343 (0.038) | 0.0001* | -0.84 (0.40) |
| Height (cm) | 0.320 (0.036) | 0.0001* | 0.352 (0.037) | 0.0001* | -0.52 (0.63) |
| BMI (kg/m2) | 0.270 (0.040) | 0.0001* | 0.289 (0.040) | 0.0001* | -0.34 (0.74) |
| Sons | MC (N=342) |  | FC (N=314) |  | Z (P) |
| Weight (kg) | 0.289 (0.050) | 0.0001* | 0.377 (0.049) | 0.0001* | -1.26 (0.21) |
| Height (cm) | 0.394 (0.047) | 0.0001* | 0.322 (0.047) | 0.0001* | 1.08 (0.23) |
| BMI (kg/m2) | 0.229 (0.056) | 0.0001* | 0.316 (0.055) | 0.0001* | -1.11 (0.27) |
| Daughters | MC (N=316) |  | FC (N=283) |  | Z (P) |
| Weight (kg) | 0.309 (0.058) | 0.0001* | 0.309 (0.060) | 0.0001* | 0 (1) |
| Height (cm) | 0.246 (0.056) | 0.0001* | 0.378 (0.058) | 0.0001* | -1.64 (0.10) |
| BMI (kg/m2) | 0.317 (0.056) | 0.0001* | 0.265 (0.057) | 0.0001* | 0.64 (0.52) |
| | 18-year<sup>\$</sup> | | | | |
| All offspring | MC (N=626)<br>beta (SE) | P-value | FC (N=568)<br>beta (SE) | P-value | Z (P) |
| Weight (kg) | 0.264 (0.040) | 0.0001* | 0.305 (0.040) | 0.0001* | -0.72 (0.47) |
| Height (cm) | 0.387 (0.032) | 0.0001* | 0.409 (0.033) | 0.0001* | -0.48 (0.63) |
| BMI (kg/m2) | 0.260 (0.042) | 0.0001* | 0.256 (0.041) | 0.0001* | 0.07 (0.95) |
| Sons | MC (N=333) |  | FC (N=307) |  | Z (P) |
| Weight (kg) | 0.252 (0.052) | 0.0001* | 0.355 (0.051) | 0.0001* | -1.41 (0.16) |
| Height (cm) | 0.391 (0.046) | 0.0001* | 0.382 (0.047) | 0.0001* | 0.14 (0.89) |
| BMI (kg/m2) | 0.245 (0.056) | 0.0001* | 0.291 (0.056) | 0.0001* | -0.58 (0.56) |
| Daughters | MC (N=293) |  | FC (N=261) |  | Z (P) |
| Weight (kg) | 0.278 (0.062) | 0.0001* | 0.247 (0.063) | 0.0001* | 0.35 (0.73) |
| Height (cm) | 0.388 (0.045) | 0.0001* | 0.444 (0.048) | 0.0001* | -0.76 (0.45) |
| BMI (kg/m2) | 0.278 (0.063) | 0.0001* | 0.219 (0.062) | 0.0001* | 0.67 (0.50) |

Values are presented as $\beta$ coefficients. \$ Parents information was considered from 12-year follow-up. Z value shows the difference between regression coefficients. $z = (b1-b2) / \sqrt{(seb1^{**2} + seb2^{**2} - cov(b1*b2))}$ ; $p = 1 - cdf.chisq(z*z,1)$ . MC: Mother-Child, FC: Father-Child. Adjusted for age, and sex.
\*significant after correcting for multiple comparisons

### Glucose and insulin indices

Fasting glucose concentrations in the offspring were positively associated with that of the mother’s as well as the father’s glucose concentrations both at 6- and 12-years. The maternal effect was stronger than the paternal with a significant parent-of-origin effect at 12-years (Table 3). The parent-of-origin effect were seen at 6- and 12-years only in sons but not daughters.

**Table 3:** Parent-of-origin effects of cardiometabolic traits. Association between offspring measures with that of each of the parents are presented as beta values and se. Differences between maternal and paternal effects are presented as Z scores and p-values.

|  | 6-year |  |  |  |  | 12-year |  |  |  |  |
| --- | --- | --- | --- | --- | --- | --- | --- | --- | --- | --- |
| All offspring | MC (N=690)<br>beta (SE) | P-value | FC (N=655)<br>beta (SE) | P-value | Z (P) | MC (N=658)<br>beta (SE) | P-value | FC (N=597)<br>beta (SE) | P-value | Z (P) |
| Fasting glucose<br>(mg/dl) | 0.321<br>(0.038) | 0.0001* | 0.231<br>(0.040) | 0.0001* | 1.63 (0.10) | 0.281<br>(0.043) | 0.0001* | 0.161<br>(0.040) | 0.0001* | 2.04 (0.04) |
| Fasting Insulin<br>(mIU/L) | -0.104<br>(0.043) | 0.016 | 0.072<br>(0.046) | 0.111 | -2.80 (0.005) * | 0.168<br>(0.044) | 0.0001* | 0.010<br>(0.045) | 0.829 | 2.51 (0.01) |
| HOMA2B | -0.035<br>(0.043) | 0.414 | 0.144<br>(0.045) | 0.001* | -2.88 (0.004) * | 0.192<br>(0.042) | 0.0001* | 0.093<br>(0.042) | 0.03 | 1.67 (0.09) |
| HOMA2S | -0.093<br>(0.043) | 0.034 | 0.068<br>(0.046) | 0.137 | -2.56 (0.01) | 0.165<br>(0.044) | 0.0001* | 0.007<br>(0.046) | 0.883 | 2.48 (0.01) |
| Triglycerides | 0.270<br>(0.043) | 0.0001* | 0.196<br>(0.041) | 0.0001* | 1.25 (0.21) | 0.276<br>(0.043) | 0.0001* | 0.310<br>(0.044) | 0.0001* | -0.55 (0.58) |
| Cholesterol | 0.320<br>(0.038) | 0.0001* | 0.169<br>(0.040) | 0.0001* | 2.72 (0.006) * | 0.427<br>(0.039) | 0.0001* | 0.254<br>(0.039) | 0.0001* | 3.14 (0.002) * |
| HDL | 0.140<br>(0.041) | 0.001* | -0.006<br>(0.041) | 0.887 | 2.52 (0.01) | 0.191<br>(0.045) | 0.0001* | 0.044<br>(0.045) | 0.333 | 2.31 (0.02) |
| Sons | 6-year |  |  |  |  | 12-year |  |  |  |  |
|  | MC (N=363) |  | FC (N=342) |  | Z (P) | MC (N=342) |  | FC (N=314) |  | Z (P) |
| Fasting glucose<br>(mg/dl) | 0.331<br>(0.048) | 0.0001* | 0.220<br>(0.044) | 0.0001 | 1.70 (0.08) | 0.230<br>(0.055) | 0.0001* | 0.075<br>(0.049) | 0.203 | 2.10 (0.03) |
| Fasting Insulin<br>(mIU/L) | -0.172<br>(0.061) | 0.005* | 0.078<br>(0.066) | 0.213 | -2.78 (0.005) * | 0.183<br>(0.068) | 0.008* | 0.060<br>(0.070) | 0.371 | 1.26 (0.20) |
| HOMA2B | -0.067<br>(0.058) | 0.266 | 0.157<br>(0.062) | 0.01 | -2.64(0.008) * | 0.191<br>(0.063) | 0.002 | 0.075<br>(0.060) | 0.216 | 1.33 (0.18) |
| HOMA2S | -0.172<br>(0.063) | 0.006* | 0.069<br>(0.066) | 0.274 | -2.64 (0.008) * | 0.178<br>(0.069) | 0.011 | 0.058<br>(0.072) | 0.394 | 1.20 (0.23) |
| Triglycerides | 0.322<br>(0.057) | 0.0001* | 0.097<br>(0.060) | 0.094 | 2.75 (0.006) * | 0.354<br>(0.058) | 0.0001* | 0.233<br>(0.060) | 0.0001* | 1.45 (0.15) |
| Cholesterol | 0.353<br>(0.054) | 0.0001* | 0.117<br>(0.054) | 0.031 | 3.09 (0.002) * | 0.464<br>(0.053) | 0.0001* | 0.233<br>(0.053) | 0.0001* | 3.08 (0.002) * |
| HDL | 0.153<br>(0.056) | 0.007* | -0.015<br>(0.057) | 0.792 | 2.10 (0.04) | 0.236<br>(0.062) | 0.0001* | -0.016<br>(0.060) | 0.796 | 2.92 (0.004) * |

| Daughters | 6-year |  |  |  |  | 12-year |  |  |  |  |
| --- | --- | --- | --- | --- | --- | --- | --- | --- | --- | --- |
|  | MC (N=327) |  | FC (N=313) |  | Z (P) | MC (N=316) |  | FC (N=283) |  | Z (P) |
| Fasting glucose<br>(mg/dl) | 0.287<br>(0.073) | 0.0001* | 0.261<br>(0.075) | 0.0001* | 0.25 (0.80) | 0.364<br>(0.073) | 0.0001 | 0.317<br>(0.068) | 0.0001 | 0.47 (0.63) |
| Fasting Insulin<br>(mIU/L) | -0.035<br>(0.061) | 0.563 | 0.050<br>(0.065) | 0.449 | -0.95 (0.31) | 0.168<br>(0.059) | 0.004 | -0.047<br>(0.059) | 0.461 | 2.62 (0.008) * |
| HOMA2B | 0.005<br>(0.065) | 0.942 | 0.123<br>(0.065) | 0.062 | -1.39 (0.16) | 0.202<br>(0.056) | 0.001 | 0.11 (0.059) | 0.073 | 1.01 (0.31) |
| HOMA2S | -0.016<br>(0.061) | 0.794 | 0.048<br>(0.066) | 0.469 | -71 (0.48) | 0.163<br>(0.058) | 0.006 | -0.049<br>(0.060) | 0.445 | 2.54 (0.01) |
| Triglycerides | 0.208<br>(0.059) | 0.0001* | 0.323<br>(0.062) | 0.0001* | -1.11 (0.27) | 0.166<br>(0.064) | 0.009 | 0.404<br>(0.063) | 0.0001 | -2.65 (0.008) * |
| Cholesterol | 0.287<br>(0.054) | 0.0001* | 0.231<br>(0.061) | 0.0001* | 0.69 (0.49) | 0.379<br>(0.058) | 0.0001 | 0.279<br>(0.057) | 0.0001 | 1.23 (0.22) |
| HDL | 0.133<br>(0.060) | 0.027 | 0.008<br>(0.060) | 0.896 | 1.47 (0.14) | 0.151<br>(0.065) | 0.021 | 0.127<br>(0.067) | 0.06 | 0.26 (0.80) |

Values are presented as $\beta$ coefficients. Z value shows the difference between regression coefficients. $z = (b1-b2) / \sqrt{se b1^{**2} + se b2^{**2} - cov(b1*b2)}$ ; $p = 1 - cdf.chisq(z*z,1)$ . MC: Mother-Child, FC: Father-Child. Adjusted for age, sex and BMI.
\*significant after correcting for multiple comparisons

A contrasting shift from 6 years to 12 years was observed for the differences between paternal and maternal effects in relation to insulin and its indices. Maternal associations were negative at 6-years and became strongly positive at 12-years, reflecting in a change of the direction of parent-of-origin effect. For fasting insulin and HOMA S, there was a significant negative maternal association at 6-years which shifted to a significant positive one at 12-years. For HOMA B, there was a stronger paternal positive effect at 6-years which changed to stronger positive maternal effect at 12 years (Table 3).

Fasting glucose showed a significant parent-of-origin effect at 6- and 12-years in sons but not daughters. For insulin and its indices, sons showed a significant negative maternal association at 6-years which shifted to a positive one at 12-years, however the parent-of-origin effects were significant only at 6-years. In the daughters, no parental associations were seen at 6-years, whereas, positive maternal associations and significant parent-of-origin effects were seen at 12-years (Table 3).

### Lipid levels

Total and HDL cholesterol levels in the offspring showed significantly stronger positive associations with mother compared to father at 6 and 12 years in all offspring as well as when analysed separately for sons. However, the parental differences were not significant in daughters at either 6 or 12-years. For triglyceride levels, sons showed strong positive maternal associations at both 6- and 12-years, however parent-of-origin effects were significant only at 6-years. In contrast, daughters showed significantly stronger paternal associations compared to maternal only at 12-years (Table 3).

## Discussion

By harnessing the potential of a birth cohort, we observed strong parent-of-origin effects on metabolic but not anthropometric traits starting from early life. Availability of serial measurements revealed changing parental influences on glucose and insulin concentrations, insulin secretion and sensitivity in the offspring. For insulin secretion, there was a transition from a predominantly paternal association in early childhood to a maternal one at pubertal age, whereas, for insulin sensitivity, a significant negative maternal association transitioned to a significantly positive one. Thus, both insulin secretion and action at pubertal age were predominantly associated with maternal phenotype. The maternal effects on lipid traits remained consistent from childhood to adolescence.

Mendelian genetics stipulates an equal contribution from each of the parents to human traits. However, parental specific influences on metabolic traits have been previously described including on beta cell response to oral glucose, insulin action in target tissues as well as lipid levels (1; 2; 11). These have been described in families of patients with type 2 diabetes and cardiovascular diseases from the Botnia and Framingham Heart Study respectively. It is of note, that both these studies investigated the POE in offspring of adult age. Since these descriptions, the developmental origins of these disorders are well established and the strongest window for ‘programming’ is thought to be periconceptional and in pregnancy (13) his suggests that parental influences should be obvious from early life. To this end, we determined the heritability and parent-of-origin effects on metabolic traits in a birth cohort that was followed up at regular intervals into young adulthood. As previously reported, these traits were robustly heritable from early childhood as when studied in adults (3; 14-16). Concordant with previous findings in support of the underlying heritable component (1; 4; 17-19), mid-parental phenotype values were significantly associated with corresponding offspring anthropometric and metabolic traits from childhood and remained so till early adulthood.

Significant parent-of-origin effects were observed for glucose, insulin and indices of insulin secretion and insulin sensitivity as well as lipid levels in the blood. Though previous studies hinted at such effects for glucose and insulin levels and their indices (1; 2; 11), our results robustly confirm these parental specific associations.

Our results of maternal effects for total and HDL cholesterol levels support the well-established previous findings (2; 11), and suggest that they manifest from childhood, albeit with some differences. A stronger correlation was reported between mother’s and daughter’s TG levels in previous studies (1; 11), however, a significant maternal effect was seen in sons at 6 years whereas a significant paternal effect was seen for daughters at 12 years in the PMNS.

Several theories have been proposed to explain the evolutionary origins of parent-of-origin effects which are a consequence of genomic imprinting. Haig and colleagues proposed the kinship theory which stipulates that imprinting is a mechanism to alter gene dosage, since there is a different effect of gene dosage on the fitness of matrilineal and patrilineal relatives (20; 21). Day and Bonduriansky put forth the sexual antagonism and offspring co-adaptation theory which states that imprinting is a mechanism to modify the resemblance of the offspring to its two parents (22; 23). Wolf and Hager’s maternal offspring coadaptation theory suggests that this kind of programming evolved to increase the probability of expressing the fitter of the two alleles at a given locus (24). The common feature of these hypotheses highlights that some processes create a selective asymmetry between the maternally and paternally inherited allele copies at a specific locus, and this causes selection to favour the differential expression of the alleles of the same locus (25). The mechanism by which this selective expression of alleles and consequent impact on the phenotype / trait is mediated by epigenetic mechanisms which are programmed in early life. Epigenetics is a link between the genes and the environment that facilitates a particular trait (26; 27); this kind of programming superimposed on top of the genetic material is by means of chemical moieties (eg DNA methylation) which can alter the way the DNA is read and expressed. Early life exposures can bring about such epigenetic reprogramming and alter development and function of organs which, in later life, can increase susceptibility to cardiometabolic disorders (27).

It has been suggested that ‘maternal constraint’ masks the paternal (genetic) influences on fetal growth (especially weight and soft tissues but not skeleton) (28; 29). Findings in the PMNS neonates hint of such an effect. Maternal phenotype significantly associated with weight and height, while, paternal association was significant only for height. For daughters, this paternal association also extended for weight. During postnatal life, maternal and paternal associations with offspring anthropometry were similar.

For insulin secretion, paternal effect in early childhood (6 years) gives way to a maternal one at 12-years. For fasting insulin and insulin sensitivity, a negative maternal effect at 6-years changed to a positive one at 12-years. The timing of the transition seemingly spans pubertal age. Metabolism and puberty are strongly interlinked; the link between nutrition and pubertal development requires the maintenance of a minimum positive energy balance, especially in females, (30-33) and undernutrition as well as overnutrition can have a significant impact on timing and progress of pubertal development and indeed fertility (30; 34). It can therefore be speculated that POE accompany pubertal changes given the increased developmental plasticity at this time period (35). Nevertheless, it remains to be seen if the shift in parental programming is a cause, consequence, or by-product linked to pubertal processes.

It is interesting to note that anthropometric measures do not show a parental specific association unlike that seen for metabolic traits. As with human traits, anthropometry is a consequence of genetics and environment, with heritability estimates increasing over time which can be partially attributed to environmental influences. We therefore suggest that this provides indirect evidence that the change in metabolic programming around pubertal age is a consequence of both genetic and epigenetic re-programming.

Our study has several limitations. The findings are observational wherein associations between parental and offspring phenotypes across trajectories of early childhood are examined and are therefore not causal. Furthermore, this study involving a single cohort and validation will be required in other similar as well as diverse populations to substantiate the findings. Nevertheless, our study is based on a large cohort with robust study power and extensive follow-up and therefore has the potential to answer novel questions in a birth cohort, as well as provide a context to findings in adult offspring from different populations. Genetic and epigenetic studies in family cohorts as well as target tissues will be very interesting to unravel mechanisms underlying these parental biases and the evolution of parental programming states.

## Data Availability

All data produced in the present study are available upon reasonable written request through a 200 word abstract to the author and owner of the data, Dr Chittaranjan Yajnik.

## Acknowledgements

We are grateful to all study participants and their family members for co-operation over many years. We thank the late Prof. DJP Barker, Prof. Caroline Fall, Dr B Coyaji, and Dr VN Rao for their support in establishing the PMNS. We thank Dr VS Padbidri and Dr L Garda, former and current Directors of Research, KEM Hospital Research Centre. We also thank the staff of the Diabetes Unit for their help in conducting the study, particularly Drs S Hirve, N Joshi, U Deshmukh, A Bavdekar as well as H Lubree, and R Ladkat, N Memane, C Joglekar, S Bagate, A Bhalerao, S Chaugule, R Dendge, T Deokar, M Gaikwad, N Gurav, S Jagtap, J Kalokhe, S Pandit, F Rajgara, D Raut, L Ramdas, M Raut, R Saswade, and V Solat. We thank Dr SS Naik, head of biochemistry, KEM Hospital for assay standardization. We are grateful to the Indian Council of Medical Research, the Department of Biotechnology, India, the Wellcome Trust and Medical Research Council, UK for their funding support. We acknowledge the support of Dr N D Deshmukh and the Zilla Parishad, Pune.We thank Assoc. Prof. Isabella Artner and Prof. Nils Wierup for critical reading of the manuscript and Senior Prof. Leif Groop for critical insights and advise on the parent-of-origin concepts.

## Author Contributions

RW analysed the data. RBP conceived the study concept and design. RW and RBP wrote manuscript. All authors took part in the interpretation of the results, commented on the manuscript, and had final responsibility for the decision to submit for publication. RBP and CSY are the guarantors of this work and, as such, had full access to all the data in the study and takes responsibility for the integrity of the data and the accuracy of the data analysis.

---

**Supplementary Table 1.** Additional description of offspring of the Pune Maternal Nutrition Study (PMNS) at birth.

| At birth | N=745 |  | N=388 |  | N=357 |  |
| --- | --- | --- | --- | --- | --- | --- |
| Birth weight (kg) | 2.7 | ±0.3 | 2.8 | ±0.3 | 2.7 | ±0.3 |
| Length (cm) | 47.3 | ±2.3 | 47.7 | ±2.5 | 47.1 | ±2.2 |
| Abdominal circumference (cm) | 28.5 | ±2.1 | 28.5 | ±2.1 | 28.4 | ±2.0 |
| Sum of skinfolds (mm) | 8.4 | ±1.7 | 8.3 | ±1.6 | 8.3 | ±1.7 |

*Values are mean ± SD*

**Supplementary Table 2:**
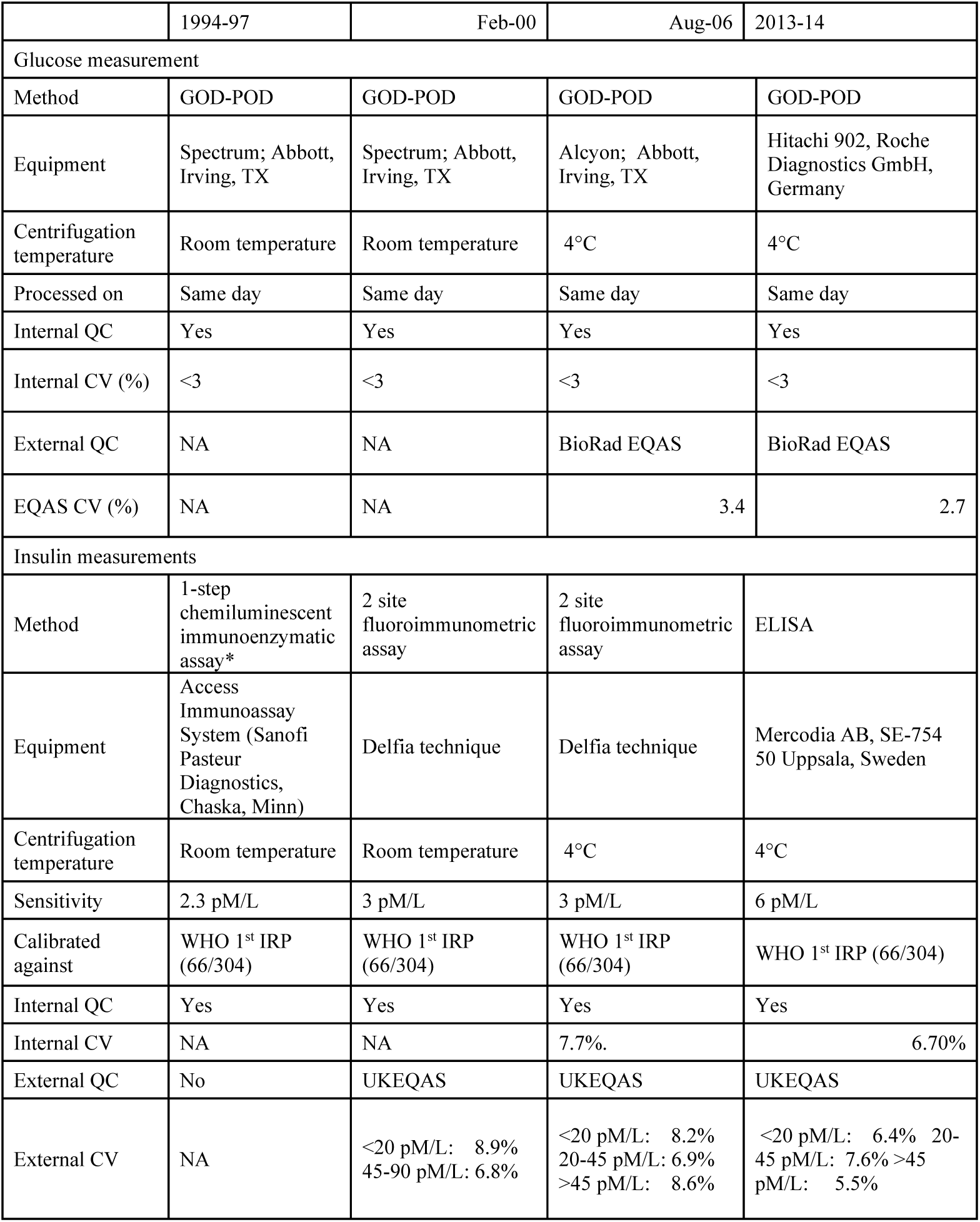
Quality assessments for glucose and insulin measurements in the Pune Maternal Nutrition Study (1993-2013).

**Supplementary Table 3.** Heritability estimates for anthropometric traits. Heritability is tested between mid-parental phenotype (predictor) and off-spring phenotype (outcome) using linear model and expressed as ß and corresponding p-values.

|  | All offspring |  |  |  |  |  |  |  |
| --- | --- | --- | --- | --- | --- | --- | --- | --- |
|  | Pre-pregnancy - At birth (N=745) |  | 6-year (N=638) |  | 12-year (N=667) |  | 12-year – 18-year (N=633) |  |
| | $\beta$ (SE) | P-value | $\beta$ | P-value | $\beta$ | P-value | $\beta$ | P-value |
| Weight (kg) | 0.187<br>(0.038) | <0.000* | 0.462<br>(0.036) | <0.000* | 0.524<br>(0.035) | <0.000* | 0.467<br>(0.037) | <0.000* |
| Height (cm) | 0.240<br>(0.037) | <0.000 | 0.509<br>(0.034) | <0.000* | 0.514<br>(0.035) | <0.000* | 0.599<br>(0.033) | <0.000* |
| BMI (kg/m<sup>2</sup>) | 0.134<br>(0.043) | 0.002* | 0.364<br>(0.037) | <0.000* | 0.440<br>(0.037) | <0.000* | 0.410<br>(0.039) | <0.000* |
| Waist $\odot$ (cm) | | | 0.239<br>(0.038) | <0.000* | | | | |
| Hip $\odot$ (cm) | | | 0.364<br>(0.036) | <0.000* | | | | |
| WHR |  |  | 0.106<br>(0.039) | <0.000* |  |  |  |  |
| Sum of skinfolds (mm) |  |  | 0.253<br>(0.038) | <0.000* |  |  |  |  |
| Total fat mass (kg) |  |  | 0.246<br>(0.038) | <0.000* |  |  |  |  |
| Total lean mass (kg) |  |  | 0.339<br>(0.037) | <0.000* |  |  |  |  |
|  | Sons |  |  |  |  |  |  |  |
|  | Pre-pregnancy- At birth (N=388) |  | 6-year (N=336) |  | 12-year (N=346) |  | 12-year – 18-year (N=339) |  |
| | $\beta$ | P-value | $\beta$ | P-value | $\beta$ | P-value | $\beta$ | P-value |
| Weight (kg) | 0.138<br>(0.049) | 0.01 | 0.480<br>(0.046) | <0.000* | 0.549<br>(0.045) | <0.000* | 0.509<br>(0.047) | <0.000* |
| Height (cm) | 0.225<br>(0.049) | <0.000* | 0.513<br>(0.045) | <0.000* | 0.556<br>(0.045) | <0.000* | 0.613<br>(0.045) | <0.000* |
| BMI (kg/m<sup>2</sup>) | 0.099<br>(0.055) | 0.087 | 0.347<br>(0.050) | <0.000* | 0.426<br>(0.051) | <0.000* | 0.425<br>(0.051) | <0.000* |
| Waist $\odot$ (cm) | | | 0.236<br>(0.049) | <0.000* | | | | |
| Hip $\odot$ (cm) | | | 0.437<br>(0.050) | <0.000* | | | | |
| WHR |  |  | 0.060<br>(0.053) | 0.262 |  |  |  |  |
| Sum of skinfolds (mm) |  |  | 0.190<br>(0.053) | <0.000* |  |  |  |  |
| Total fat mass (kg) |  |  | 0.225<br>(0.050) | <0.000* |  |  |  |  |
| Total lean mass (kg) |  |  | 0.314<br>(0.049) | <0.000* |  |  |  |  |
|  | Daughters |  |  |  |  |  |  |  |
|  | Pre-pregnancy- At birth (N=357) |  | 6-year (N=302) |  | 12-year (N=321) |  | 12-year – 18-year (N=294) |  |
| | $\beta$ | P-value | $\beta$ | P-value | $\beta$ | P-value | $\beta$ | P-value |
| Weight (kg) | 0.253<br>(0.059) | <0.000* | 0.441<br>(0.056) | <0.000* | 0.500<br>(0.056) | <0.000* | 0.415<br>(0.059) | <0.000* |
| Height (cm) | 0.260<br>(0.056) | <0.000* | 0.505<br>(0.052) | <0.000* | 0.466<br>(0.057) | <0.000* | 0.579<br>(0.048) | <0.000* |
| BMI<br>(kg/m<sup>2</sup>) | 0.178<br>(0.068) | 0.005 | 0.380<br>(0.055) | <0.000* | 0.461<br>(0.055) | <0.000* | 0.398<br>(0.059) | <0.000* |
| Waist Ø<br>(cm) |  |  | 0.244<br>(0.059) | <0.000* |  |  |  |  |
| Hip Ø (cm) |  |  | 0.291<br>(0.053) | <0.000* |  |  |  |  |
| WHR |  |  | 0.158<br>(0.056) | 0.005* |  |  |  |  |
| Sum of<br>skinfolds<br>(mm) |  |  | 0.323<br>(0.054) | <0.000* |  |  |  |  |
| Total fat<br>mass (kg) |  |  | 0.271<br>(0.057) | <0.000* |  |  |  |  |
| Total lean<br>mass (kg) |  |  | 0.371<br>(0.051) | <0.000* |  |  |  |  |

**Supplementary Table 4:** Heritability estimate for glycemic traits. Heritability is tested between mid-parental phenotype (predictor) and off-spring phenotype (outcome) using linear model and expressed as ß and corresponding p-values.

|  | All offspring |  |  |  |
| --- | --- | --- | --- | --- |
|  | 6-year (N=638) |  | 12-year (N=667) |  |
| | $\beta$ | P-value | $\beta$ | P-value |
| Fasting glucose (mg/dl) | 0.409 (0.036) | <0.000* | 0.307 (0.040) | <0.000* |
| Fasting Insulin (mIU/L) | 0.026 (0.039) | 0.503 | 0.198 (0.041) | <0.000* |
| HOMA2B | 0.104 (0.039) | 0.008* | 0.241 (0.040) | <0.000* |
| HOMA2S | -0.005 (0.039) | 0.906 | 0.202 (0.041) | <0.000* |
| Triglycerides | 0.301 (0.037) | <0.000* | 0.378 (0.039) | <0.000* |
| Cholesterol | 0.373 (0.036) | <0.000* | 0.449 (0.037) | <0.000* |
| HDL | 0.108 (0.039) | 0.006* | 0.169 (0.041) | <0.000* |
|  | Sons |  |  |  |
|  | 6-year (N=336) |  | 12-year (N=346) |  |
| | $\beta$ | P-value | $\beta$ | P-value |
| Fasting glucose (mg/dl) | 0.383 (0.044) | <0.000* | 0.188 (0.048) | 0.001* |
| Fasting Insulin (mIU/L) | 0.004 (0.056) | 0.938 | 0.179 (0.056) | 0.002* |
| HOMA2B | 0.110 (0.055) | 0.043 | 0.206 (0.055) | <0.000* |
| HOMA2S | -0.045 (0.057) | 0.409 | 0.201 (0.060) | <0.000* |
| Triglycerides | 0.249 (0.053) | <0.000* | 0.339 (0.054) | <0.000* |
| Cholesterol | 0.338 (0.055) | <0.000* | 0.419 (0.052) | <0.000* |
| HDL | 0.114 (0.053) | 0.034 | 0.182 (0.056) | 0.001* |
|  | Daughters |  |  |  |
|  | 6-year (N=302) |  | 12-year (N=321) |  |
| | $\beta$ | P-value | $\beta$ | P-value |
| Fasting glucose (mg/dl) | 0.464 (0.062) | <0.000* | 0.536 (0.067) | <0.000* |
| Fasting Insulin (mIU/L) | 0.049 (0.055) | 0.384 | 0.224 (0.060) | <0.000* |
| HOMA2B | 0.100 (0.056) | 0.077 | 0.285 (0.058) | <0.000* |
| HOMA2S | 0.037 (0.055) | 0.502 | 0.207 (0.057) | 0.001* |
| Triglycerides | 0.359 (0.053) | <0.000* | 0.423 (0.055) | <0.000* |
| Cholesterol | 0.411 (0.052) | <0.000* | 0.482 (0.054) | <0.000* |
| HDL | 0.101 (0.057) | 0.074 | 0.155 (0.060) | 0.01 |

**Supplementary Table 5.** Parent-of-origin test for offspring anthropometry for additional phenotypes at 6-years of age. Association between offspring anthropometry measures with that of each of the parents are presented as beta values and se. Differences between maternal and paternal effects are presented as Z scores and p-values.

|  | All offspring |  |  |  |  |
| --- | --- | --- | --- | --- | --- |
|  | MC (N=690) | P-value | FC (N=655) | P-value | Z (P) |
| Waist O (cm) | 0.192 (0.039) | 0.0001 | 0.142 (0.039) | 0.0001 | 0.91 (0.36) |
| Hip O (cm) | 0.258 (0.040) | 0.0001 | 0.261 (0.038) | 0.0001 | -0.05 (0.96) |
| WHR | 0.139 (0.040) | 0.0001 | 0.037 (0.040) | 0.348 | 1.80 (0.07) |
| Sum of skinfolds (mm) | 0.123 (0.039) | 0.002 | 0.211 (0.040) | 0.0001 | -1.58 (0.12) |
| Total fat mass (kg) | 0.130 (0.040) | 0.001 | 0.187 (0.040) | 0.0001 | -1.01 (0.31) |
| Total lean mass (kg) | 0.251 (0.037) | 0.0001 | 0.262 (0.038) | 0.0001 | -0.21 (0.84) |

|  | Sons |  |  |  |  |
| --- | --- | --- | --- | --- | --- |
|  | MC (N=363) | P-value | FC (N=342) | P-value | Z (P) |
| Waist O (cm) | 0.190 (0.051) | 0.001 | 0.144 (0.052) | 0.009 | 0.63 (0.53) |
| Hip O (cm) | 0.271 (0.048) | 0.0001 | 0.273 (0.051) | 0.0001 | -0.03 (0.98) |
| WHR | 0.099 (0.052) | 0.073 | 0.041 (0.053) | 0.454 | 0.78 (0.43) |
| Sum of skinfolds (mm) | 0.123 (0.054) | 0.028 | 0.139 (0.056) | 0.014 | -0.21 (0.84) |
| Total fat mass (kg) | 0.082 (0.054) | 0.154 | 0.184 (0.057) | 0.001 | -1.30 (0.19) |
| Total lean mass (kg) | 0.229 (0.050) | 0.0001 | 0.264 (0.049) | 0.0001 | -0.50 (0.62) |

|  | Daughters |  |  |  |  |
| --- | --- | --- | --- | --- | --- |
|  | MC (N=327) | P-value | FC (N=313) | P-value | Z (P) |
| Waist O (cm) | 0.197 (0.061) | 0.001 | 0.139 (0.060) | 0.015 | 0.68 (0.50) |
| Hip O (cm) | 0.246 (0.069) | 0.0001 | 0.245 (0.058) | 0.0001 | -0.01 (0.99) |
| WHR | 0.196 (0.066) | 0.001 | 0.035 (0.060) | 0.536 | 1.81 (0.07) |
| Sum of skinfolds (mm) | 0.127 (0.057) | 0.024 | 0.292 (0.057) | 0.0001 | -2.05 (0.04) |
| Total fat mass (kg) | 0.185 (0.060) | 0.001 | 0.201 (0.058) | 0.0001 | -0.19 (0.85) |
| Total lean mass (kg) | 0.276 (0.056) | 0.0001 | 0.260 (0.059) | 0.0001 | 0.20 (0.84) |

Values are presented as $\beta$ coefficients. Z value shows the difference between regression coefficients. $z = (b1-b2) / \sqrt{seb1^{**2} + seb2^{**2} - cov(b1*b2)}$ ; $p = 1 - cdf.chisq(z*z,1)$ . MC: Mother-Child, FC: Father-Child. Adjusted for age, sex and BMI.

